# Autoantigen profiling reveals a shared post-COVID signature in fully recovered and Long COVID patients

**DOI:** 10.1101/2023.02.06.23285532

**Authors:** Aaron Bodansky, Chung-Yu Wang, Aditi Saxena, Anthea Mitchell, Saki Takahashi, Khamal Anglin, Beatrice Huang, Rebecca Hoh, Scott Lu, Sarah A. Goldberg, Justin Romero, Brandon Tran, Raushun Kirtikar, Halle Grebe, Matthew So, Bryan Greenhouse, Matthew S. Durstenfeld, Priscilla Y. Hsue, Joanna Hellmuth, J. Daniel Kelly, Jeffrey N. Martin, Mark S. Anderson, Steven G. Deeks, Timothy J. Henrich, Joseph L. DeRisi, Michael J. Peluso

**Author notes:** Corresponding Author: Michael Peluso, MD, Division of HIV, Infectious Diseases, and Global Medicine University of California, San Francisco, San Francisco, CA, USA.

## Abstract

Some individuals do not return to baseline health following SARS-CoV-2 infection, leading to a condition known as Long COVID. The underlying pathophysiology of Long COVID remains unknown. Given that autoantibodies have been found to play a role in severity of COVID infection and certain other post-COVID sequelae, their potential role in Long COVID is important to investigate. Here we apply a well-established, unbiased, proteome-wide autoantibody detection technology (PhIP-Seq) to a robustly phenotyped cohort of 121 individuals with Long COVID, 64 individuals with prior COVID-19 who reported full recovery, and 57 pre-COVID controls. While a distinct autoreactive signature was detected which separates individuals with prior COVID infection from those never exposed to COVID, we did not detect patterns of autoreactivity that separate individuals with Long COVID relative to individuals fully recovered from SARS-CoV-2 infection. These data suggest that there are robust alterations in autoreactive antibody profiles due to infection; however, no association of autoreactive antibodies and Long COVID was apparent by this assay.

## BACKGROUND

Some individuals do not fully return to baseline health following SARS-CoV-2 infection and experience ongoing morbidity following the acute phase of COVID-19 (1, 2). There is now intense interest in determining the underlying mechanisms of Long COVID, one type of post-acute sequelae of SARS-CoV-2 infection (PASC) characterized by symptoms that newly developed or worsened following infection that cannot be clearly attributed to another cause (3). Immune dysregulation, including the generation of antibodies against self antigens, has been suggested as one potential driver of Long COVID that warrants further investigation (4, 5).

Acute SARS-CoV-2 infection is associated with the generation of autoreactive antibodies, particularly among individuals with severe disease requiring hospitalization (6–8). For example, one study of 147 individuals hospitalized with COVID-19 found that autoantibodies associated with connective tissue diseases and anti-cytokine antibodies were identified in 50% of samples, and tracked with the humoral response to SARS-CoV-2 infection (7). Furthermore, multiple studies have described the contribution of likely pre-existing anti-interferon antibodies to vaccine breakthrough infections and severe manifestations of COVID-19, including death (9–12).

Although much work has been done exploring the potential virologic and immunologic factors driving Long COVID, the evaluation of autoantibodies in this condition has been more limited. Measurement of anti-nuclear antibodies on standard clinical tests has yielded mixed findings, with some studies identifying a high prevalence of antinuclear antibodies (ANAs) among those with Long COVID (13–15) and other studies finding low prevalence consistent with ANA positivity in the general population (16–18). Although perturbations in interferon signaling pathways have been suggested as one potential mechanism of Long COVID (4, 15), anti-interferon antibodies have not been identified in most individuals outside of those who had severe acute infection (16). This is consistent with other post-acute sequelae of COVID such as multisystem inflammatory syndrome in children (MIS-C), which also has no association with anti-interferon antibodies (19).

The identification of previously reported autoantibodies can be performed using targeted assays. However, identifying the full range of novel autoreactive antibodies, whether they are pathological or not, requires technologies capable of high-throughput, unbiased, proteome-wide screens. In this study, we screened a cohort of individuals with prior SARS-CoV-2 infection, many of whom met clinical criteria for Long COVID, to determine whether a consistent pattern of autoreactivity could be identified. This same technology has been previously utilized to discover novel autoantibodies in a wide range of disease contexts (20–23).

## RESULTS

### Distinct set of autoreactive antibodies in individuals with prior SARS-CoV-2 infection

We employed a previously published proteome-wide approach using a T7 phage-display assay with immunoprecipitation and next-generation sequencing (PhiP-Seq) (20–24). We tested sera from 185 otherwise healthy individuals with prior SARS-CoV-2 infection in parallel to sera from 57 otherwise healthy individuals collected prior to the known existence of COVID-19 (pre-COVID). Using an unbiased analysis, we identified a distinct pattern of autoreactivity which effectively classifies individuals with prior SARS-CoV-2 infection from individuals not yet exposed to the virus with a logistic regression AUC of 0.90 (Figure 1A). The protein targets from which these enriched immunoprecipitated peptides were derived are widely varied and lack any apparent shared biological functions or cell type (Figure 1B). Among the identified targets, a single peptide derived from ARHGAP31 (Figure 2A) displayed the greatest amount of enrichment with 22% of individuals with prior SARS-CoV-2 infection yielding enrichment greater than 6 standard deviations of the mean of the pre-COVID controls. No difference in enrichment was observed for those diagnosed with Long COVID with respect to Post-COVID (Figure 2B).

**Figure 1:**
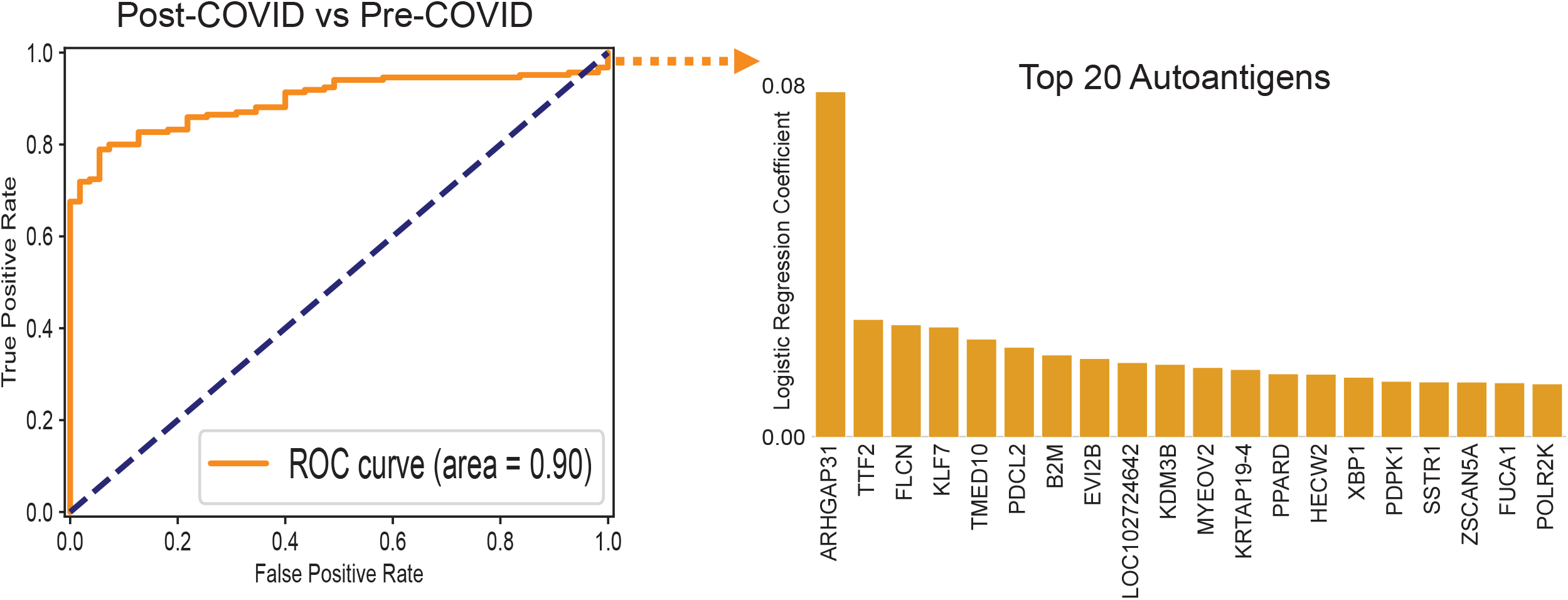
PhIP-Seq autoreactivities distinguish post-COVID sera from pre-COVID controls. Logistic regression comparing PhIP-Seq autoreactivities in all individuals with prior COVID infection compared to pre-COVID controls. Barplot showing autoreactivities with the top 20 logistic regression coefficients.

**Figure 2:**
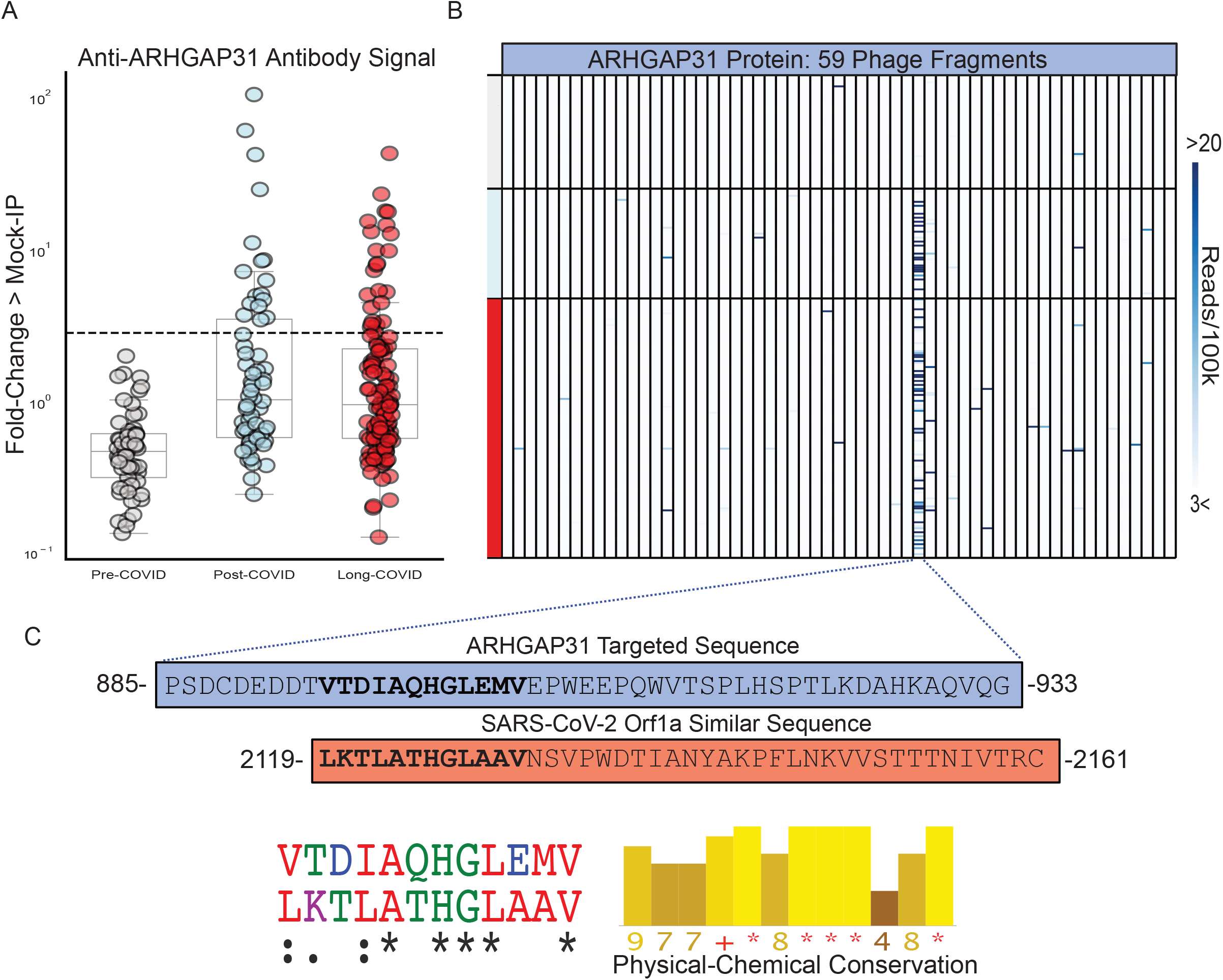
Post-COVID anti-ARHGAP31 autoreactivities target specific region with similarity to SARS-CoV-2. (A) Stripplots showing distribution of ARHGAP31 autoreactivities in Long COVID, individuals with prior COVID infection but without Long COVID, and pre-COVID controls. Dotted line at 6 standard deviations above mean of pre-COVID controls. (B) Distribution of anti-ARHGAP31 autoreactivity signal within ARHGAP31 full length protein. One specific fragment is targeted. (C) Amino-acid sequence of the autoreactive region of ARHGAP31 and amino-acid sequence of a region of SARS-CoV-2 Orf1a with similarity. Shown below is the multiple sequence alignment(ClustalOmega; asterisk=identical amino acid; colon=strongly similar properties with Gonnet PAM 250 matrix score >0.5; period=weakly similar with Gonnet PAM 250 matrix score between 0 and 0,5) and strong physical-chemical conservation (JalView; amino acid physical-chemical conservation scored on a scale of 1-11, Asterisk=score of 11 and identical amino acid, Plus=10, all properties conserved).

To investigate whether the ARHGAP31 peptide enrichment could be the result of cross-reactivity with antibodies directed against SARS-CoV-2 proteins, we performed a multiple sequence alignment of this 49-amino acid fragment against the full SARS-CoV-2 proteome. A region within the SARS-CoV2 Orf1a polyprotein was identified with considerable physico-chemical similarity to a portion of the autoreactive fragment in ARHGAP31(Jalview Version 2;Figure 2C), supporting the notion that the observed human peptidome peptide enrichments in post-COVID samples are being driven by anti-SARS-CoV-2 antibodies.

ARHGAP31 is most highly expressed in neuronal cells, Langerhans cells, and endothelial cells (Human Protein Atlas, proteinatlas.org). We were unable to identify any clinical differences between individuals with and without ARHGAP31 autoreactivity, such as differences in frequency of neurologic symptoms.

### Post-COVID autoreactivities are not enriched in individuals with Long COVID

To assess whether autoreactive antibodies present in individuals following SARS-CoV-2 infection are associated with Long COVID, we compared the distribution of the enriched post-COVID peptides amongst the 121 individuals with Long COVID and the 64 individuals with prior SARS-CoV-2 infection but without Long COVID (Convalescent COVID). The 20 most enriched proteins with at least 5-fold greater than background (defined as fold-change over mock-IP with protein A/G beads) were compared (Figure 3). Seventeen of the 20 enriched proteins were present in both Long COVID and Convalescent COVID, and none of these enrichments was observed in any of the pre-COVID controls. Overall, there was no significant differences in enrichment between Long COVID and Convalescent COVID. Peptides derived from three proteins, TMED10, FUCA1, and POL2RK, were only observed in Long COVID, however these were infrequently observed.

**Figure 3:**
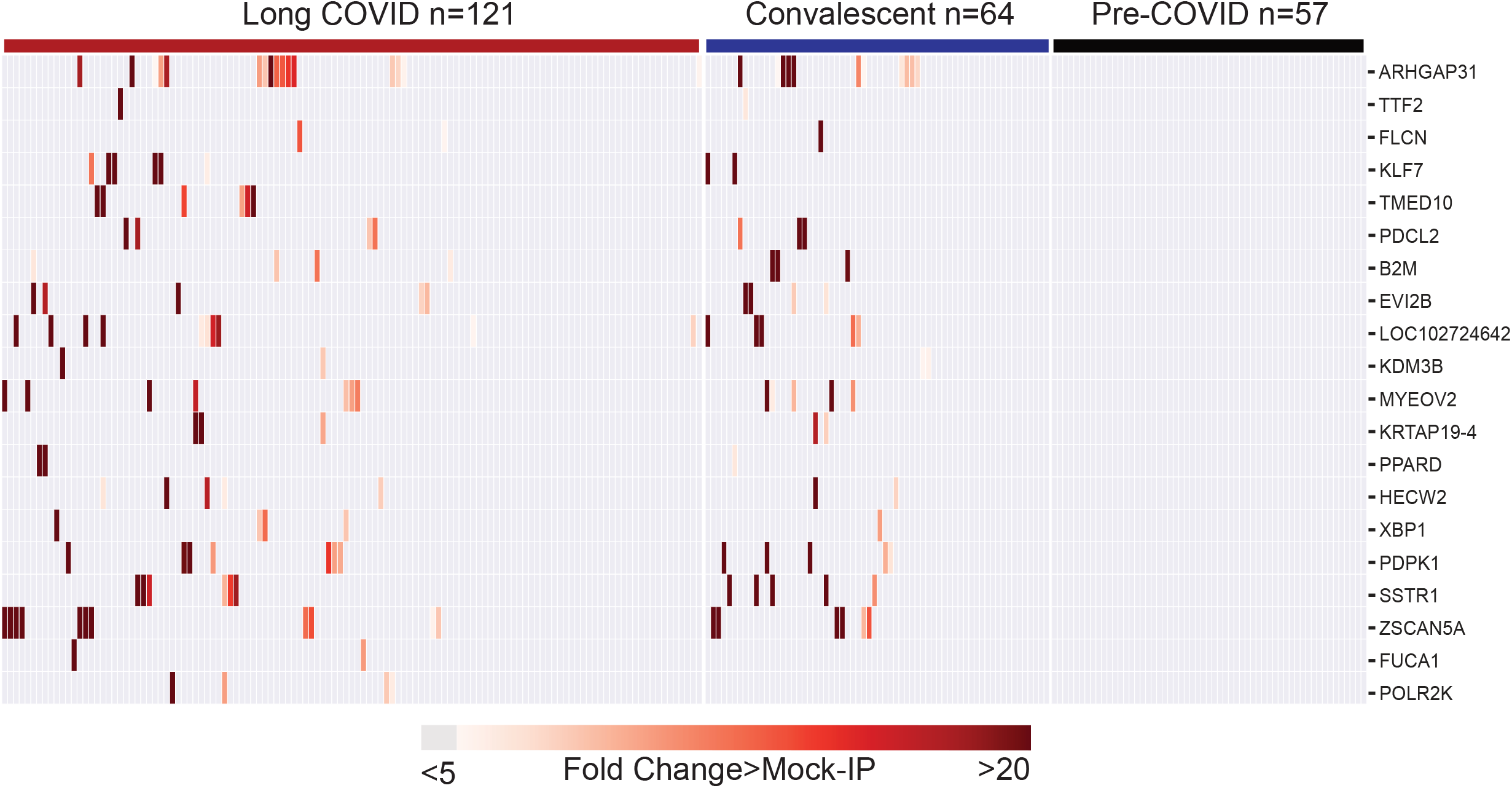
Post-COVID autoreactivities are similarly distributed among Long COVID and controls. Hierarchically clustered (Pearson) heatmaps showing the PhIP-Seq enrichment for the top 20 autoreactivities ranked by logistic regression coefficient in each Long COVID patient, each COVID convalescent patient, and each pre-COVID control.

Subgroup analyses were performed to determine whether any of these observed autoreactivities were enriched in particular symptom-defined phenotypes of Long COVID. These include cardiopulmonary (cough, shortness of breath, chest pain, palpitations, and fainting), central neurologic symptoms (problems with vision, headache, difficulty with concentration or memory, dizziness, and difficulty with balance), any neurologic symptom (problems with vision, headache, difficulty with concentration or memory, dizziness, difficulty with balance, trouble with smell or taste, phantosmia, or paresthesia), gastrointestinal (diarrhea, constipation, nausea, vomiting, loss of appetite, and abdominal pain), musculoskeletal (back pain, muscle pain, pain in the arms, legs, or joints), and upper respiratory (runny nose and sore throat).

The peptide enrichments from individuals with particular Long COVID phenotypes were compared to the enrichments from individuals with Convalescent COVID (no Long COVID symptoms) using a one-sided Kolmogorov-Smirnov test. None of the top 20 autoantibodies were enriched in severe Long COVID, and only 3 autoantibodies were statistically increased in any phenotype: TTF2 and KDM3B in those with cardiopulmonary symptoms and FUCA1 in those with upper respiratory symptoms (Figure 4A). However, using a strict cutoff of 6 standard deviations above the pre-COVID controls to determine positivity, none of these autoantibodies was phenotype specific. By looking at the distribution of these antibodies in Long COVID patients with given sub-phenotype relative to the remaining Long COVID patients and all convalescent COVID patients, it was apparent that the statistical significance in a particular phenotype was driven by either a single individual with extremely high autoantibody signal in the case of KDM3B and TTF2, or higher group signal but not meeting the positive threshold cutoff in the case of FUCA1 (Figure 4B).

**Figure 4:**
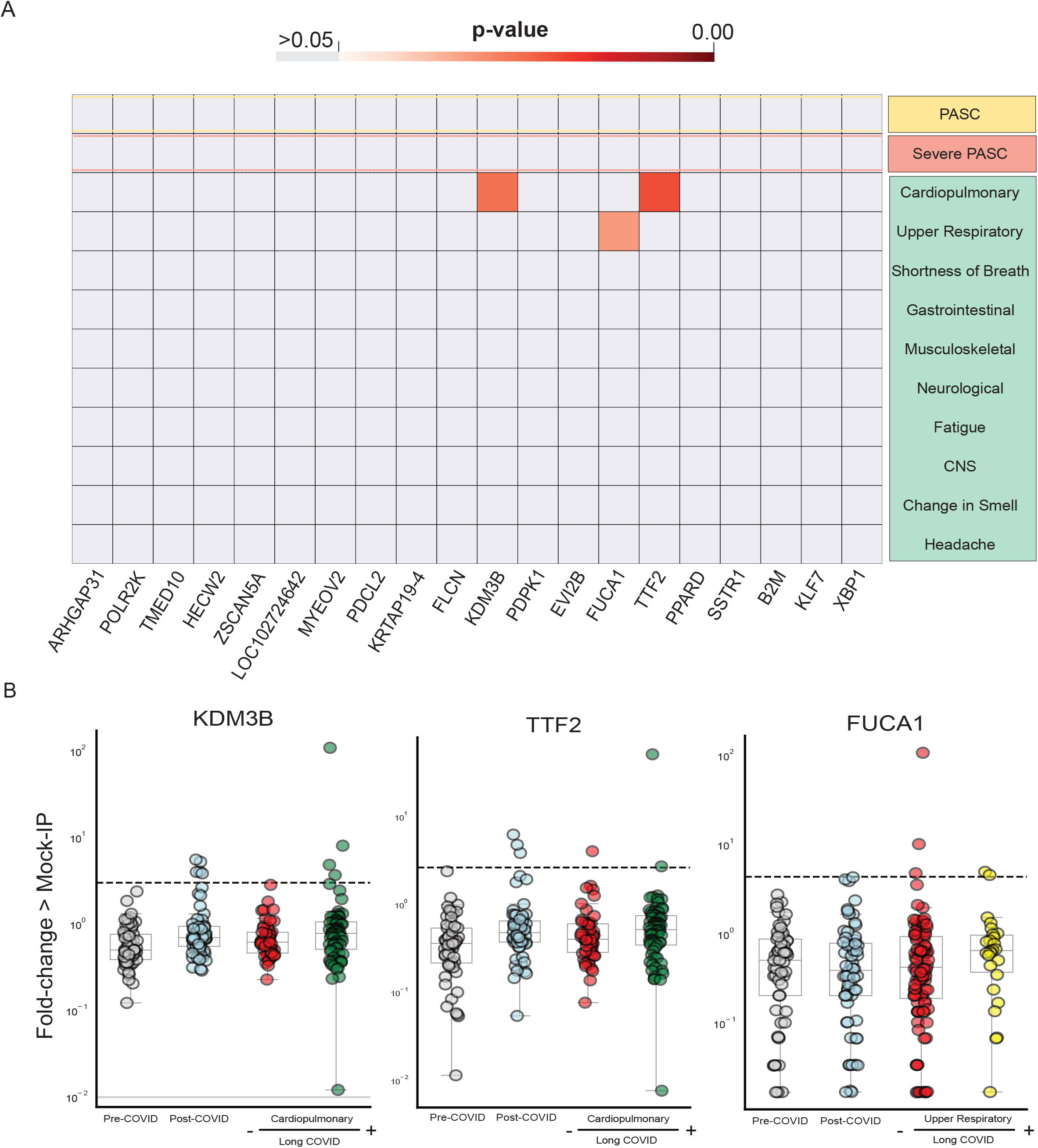
Few significantly increased autoreactivities in Long COVID symptom phenotypes. Heatmap with p-values (Kolmogorov-Smirnov testing) of differences in autoantigen enrichment for all individuals with prior COVID infection with and without additional clinical factors. Top-row compares those with and without Long COVID. Lower rows show subcategories of Long COVID. (B) Stripplots showing the three autoantibodies with statistically significant enrichment in a post-COVID clinical phenotype. Dotted-lines show 6 standard-deviations above the mean of pre-COVID signal.

### Absence of Long COVID specific autoreactivities

In addition to analyzing the distribution of post-COVID autoreactive peptide enrichments, we also performed additional analyses to detect those enrichments which might be present only in Long COVID or particular subcategories of Long COVID. To feature-weight enriched peptides, we applied logistic regressions to the degree of enrichment for each individual with Long COVID and each individual Long COVID symptom phenotype versus enrichment from convalescent COVID patients who did not have the particular symptoms. We included all previously discussed symptom phenotypes, as well as individuals with difficulty concentrating, worsened quality of life, new depression, and generalized anxiety disorder. We were unable to identify a set of enriched proteins specific to any of these groups of patients which could effectively distinguish the cohort from controls, as the best receiver operating characteristic (ROC) area under the curve (AUC) was 0.67, and the mean AUC was 0.44 (Supplement 1).

## DISCUSSION

Autoimmunity has been proposed as one potential mechanism driving Long COVID. We applied an unbiased, proteome-wide, validated approach to assess associations between antibody autoreactivity and clinical phenotype. A clear and robust difference in autoreactivity was detected between those infected with SARS-CoV-2 and pre-COVID controls. This difference was constituted by peptides from diverse and varied proteins, most of which are intracellular, suggesting that the origin of the differential enrichment is due to cross-reactivity with SARS-CoV-2 directed antibodies in those that were exposed. A sequence comparison between a peptide from the most enriched protein, ARHGAP31, and Orf1a of SARS-CoV-2 supports this notion, but orthogonal validation through fine-scale epitope mapping and antibody cloning would be required to demonstrate this conclusively. While the clinical significance of incidental autoreactivity due to the humoral immune response to SARS-CoV-2 remains largely unknown, prior studies have identified cross reactive autoantibodies in severe sequelae of SARS-CoV-2 infection, including those that develop severe neurological symptoms (Song et al., 2021). Understanding the clinical consequences of SARS-CoV-2 driven autoreactivity deserves further attention, perhaps through long-term longitudinal studies.

We found no association to support the hypothesis that autoreactivity, as detected in this assay, contributes to Long COVID. Despite numerous successes, PhIP-seq possesses a number of limitations. Because the T7 phage displayed peptides are only 49 amino acids, the assay inherently detects mostly linear epitopes. Therefore, complex conformational, post translationally modified, or multimeric protein configurations are not predicted to be detectable by this assay, and thus these results do not completely rule out autoimmune interactions in Long COVID that are beyond the scope of PhIP-seq.

In addition, our classification of Long COVID and its symptom phenotypes was based exclusively on participant self-report of symptoms. It is possible that future analyses in more homogenous cohorts, particularly those with objectively measured physiologic perturbations now associated with certain Long COVID phenotypes (e.g., postural orthostasis/tachycardia, neurocognitive function deficits, abnormalities on cardiopulmonary exercise testing) may yet reveal a role of autoantibodies in at least a subset of individuals experiencing Long COVID.

Long COVID remains a complex clinical entity. Its causes are likely multifactorial, and there is growing consensus that different phenotypes are driven by different pathophysiologic mechanisms (2, 3). Additional work characterizing SARS-CoV-2 specific and autoreactive immune responses in large, well-characterized cohorts over time, during both the acute and post-acute phases of the illness, will be necessary to delineate the biology of Long COVID and other post-acute sequelae of SARS-CoV-2 infection, and to lead to the development of potential interventions to treat the millions of individuals currently affected by this condition.

## METHODS

### Study participants and measurements

Participants were volunteers in the UCSF Long-term Impact of Infection with Novel Coronavirus (LIINC) study (NCT04362150). The details of study design and measurement have been reported previously (25). For the current analysis, we included 185 individuals with a history of nucleic acid-confirmed SARS-CoV-2 infection who had a plasma sample collected between 60 and 240 days following initial symptom onset or, if asymptomatic, first positive SARS-CoV-2 test (Supplemental Table 1). Long COVID was defined using study instruments, which have been described in detail elsewhere (25). Briefly, each participant was queried regarding the presence, severity, and duration of 32 physical and mental health symptoms and quality of life. Details of their medical history and COVID-19 infection and vaccination and treatment history were also recorded.

Symptoms that pre-dated SARS-CoV-2 infection and that were not changed following infection, as well as those obviously attributed to another cause (e.g. ankle fracture), were not considered to represent Long COVID. All samples included were collected prior to the subject ever receiving a SARS-CoV-2 vaccine.

Because of challenges in objectively defining Long COVID and to thoroughly explore the data in an unbiased manner, we utilized a number of predefined case definitions in our analysis. We constructed case definitions based on symptom presentation and quality-of-life (QOL) responses. Symptoms case definitions include: the presence of any new or worsening symptoms since SARS-CoV-2 infection (Long COVID); presence of 5 or more symptoms (severe Long COVID); specific symptom groups according to organ system involvement or phenotypic cluster; and individual symptoms when at least 25 individuals experienced the symptom. For individual and grouped symptom outcomes, we developed 3 potential comparisons between the symptomatic group and (1) all individuals who reported absence of the symptoms of interest, regardless of Long COVID status (2) only individuals who were consistently asymptomatic and (3) individuals with Long COVID but not the symptoms of interest. Severe Long COVID was compared only to those without any reported symptoms.

We defined 6 groups of symptoms (symptom phenotypes) based on either organ system cluster. These include cardiopulmonary (cough, shortness of breath, chest pain, palpitations, and fainting), CNS-specific (problems with vision, headache, difficulty with concentration or memory, dizziness, and difficulty with balance), any neurologic symptom (problems with vision, headache, difficulty with concentration or memory, dizziness, difficulty with balance, trouble with smell or taste, phantosmia, or paresthesia), gastrointestinal (diarrhea, constipation, nausea, vomiting, loss of appetite, and abdominal pain), musculoskeletal (back pain, muscle pain, pain in the arms, legs, or joints), and upper respiratory (runny nose and sore throat).

Quality-of-life was assessed using the EuroQol-5D (EQ-5D), Patient Health Questionnaire depression scale (PHQ-8), and Generalized Anxiety Disorder scale (GAD-7). Individuals with Long COVID and the lowest overall quality-of-life (QOL) score measured via the visual analogue scale of the EQ-5D were compared to individuals with the highest overall QOL scores among those with and without Long COVID. Individuals with responses categorized as “moderate depression” on the PHQ-8 (score higher than 10) and “moderate anxiety” on the GAD-7 (score higher than 9) were compared to all participants with scores indicating less severe classifications than “moderate depression” and “moderate anxiety” in the following groups: (1) all participants regardless of Long COVID status (2) all participant without Long COVID and (3) all participants with Long COVID.

In addition, we recently demonstrated associations between Long COVID and other chronic latent viral infections, including serologic evidence suggesting recent Epstein-Barr virus (EBV) reactivation (26). For this reason, we also used binary variables to create groups indicating the presence of this condition.

### Biospecimen collection

At each visit, whole blood was collected in EDTA tubes. Plasma was isolated and stored in -80F until the time of analysis.

### Phage Immunoprecipitation and Sequencing (PhIP-Seq)

PhIP-Seq was performed following our previously published vacuum-based PhIP-Seq protocol (23) (https://www.protocols.io/view/scaled-high-throughput-vacuum-phip-protocol-ewov1459kvr2/v1).

### PhIP-Seq analysis

All analysis (except when specifically stated otherwise) was performed at the gene-level, in which all reads for all peptides mapping to the same gene were summed, and 0.5 reads were added to each gene to allow inclusion of genes with zero reads in mathematical analyses. Within each individual sample, reads were normalized by converting to the percentage of total reads. To normalize each sample against background non-specific binding, a fold-change(FC) over mock-IP was calculated by dividing the sample read percentage for each gene by the mean read-percentage of the same gene for the AG bead only controls. This FC signal was then used for side-by-side comparison between samples and cohorts. Samples which had a FC of 5 or greater were considered enriched for an antibody, and samples with a FC of 6 standard deviations above the mean of pre-COVID controls were considered positive for an autoantibody. FC values were also used to calculate z-scores for each disease category sample by using each respective control (as specified in figures and results), and for each control sample by using all remaining controls. These z-scores were used for the logistic-regression feature weighting. In the case peptide-level analysis, raw reads were normalized by calculating the number of reads per 100,000 reads.

### Statistical methods

All statistical analysis was performed in Python using the Scipy Stats package. A two-way Kolmogorov-Smirnoff(KS) test was used for comparisons of FC PhIP-Seq data between groups of samples, except in the case of specifically looking for those genes with increased signal only in the disease-cohort in which a one-way KS test was employed. The logistic regression machine-learning classifiers were performed using our recently described methods (23). Utilizing the Scikit-learn package, logistic regression classifiers were applied to z-scored PhIP-Seq values from individuals with a designated disease category versus the designated control. A liblinear solver was used with L1 regularization, and the model was evaluated using a five-fold cross-validation (4 of the 5 for training, 1 of the 5 for testing).

### Informed consent

Participants provided written informed consent. The study was approved by the UCSF Institutional Review Board.

## Data Availability

All data produced in the present study are available upon reasonable request to the authors.

## FOOTNOTES

## Acknowledgements

We are grateful to the study participants and their medical providers. We acknowledge current and former LIINC clinical study team members Tamara Abualhsan, Andrea Alvarez, Khamal Anglin, Urania Argueta, Mireya Arreguin, Kofi Asare, Melissa Buitrago, Monika Deswal, Nicole DelCastillo, Emily Fehrman, Halle Grebe, Heather Hartig, Yanel Hernandez, Rebecca Hoh, Beatrice Huang, Marian Kerbleski, Raushun Kirtikar, James Lombardo, Monica Lopez, Michael Luna, Sadie Munter, Lynn Ngo, Enrique Martinez Ortiz, Antonio Rodriguez, Justin Romero, Dylan Ryder, Ruth Diaz Sanchez, Matthew So, Celina Chang Song, Viva Tai, Alex Tang, Cassandra Thanh, Fatima Ticas, Leonel Torres, Brandon Tran, Daisy Valdivieso, Deepshika Varma, and Meghann Williams; and LIINC laboratory team members Amanda Buck, Tyler-Marie Deveau, Joanna Donatelli, Jill Hakim, Nikita Iyer, Owen Janson, Brian LaFranchi, Christopher Nixon, Isaac Thomas, and Keirstinne Turcios. We thank Jessica Chen, Aidan Donovan, Carrie Forman, and Rania Ibrahim for assistance with data entry and review. We thank the UCSF AIDS Specimen Bank for processing specimens and maintaining the LIINC biospecimen repository. We are grateful to Elnaz Eilkhani and Monika Deswal for regulatory support.

## Author Contributions

AB, ST, BG, JDT, JNM, SGD, TJH, JLD, and MJP designed the study. JDT, JNM, SGD, TJH, and MJP designed and oversaw the LIINC cohort, with major contributions from BG, MSD, PYH, and JH. KA, BH, and RH managed cohort operations. Clinical data were collected by JR, BT, RK, HG, and MS. SL and SG managed the data and developed the clinical dataset. ST, SL, SG, and MJP selected participants for inclusion in this study. Biospecimens were processed in the laboratories of BG and TJH. CW, AS, and AM performed the measurements in the laboratories of MSA and JLD. AB performed the data analysis. AB, TJH, JLD, and MJP wrote the first draft of the paper, with input from all other authors. All authors edited the manuscript and approved the final version.

## Funding

This work was supported by NIH/NIAID 3R01AI141003-03S1 and NIH/NIAID R01AI158013 (to M Gandhi and M Spinelli), Pediatric Scientist Development Program and the Eunice Kennedy Shriver National Institute of Child Health and Human Development (K12-HD000850 to A. Bodansky), and Chan Zuckerberg Biohub support for J. DeRisi.

## Conflicts of Interest

TJH reports grants from Merck & Co, Gilead Sciences, and Bristol-Myers Squibb, and has provided consulting for Roche, outside the submitted work. SGD reports grants and/or personal fees from Gilead Sciences, Merck & Co., Viiv, AbbVie, Eli Lilly, ByroLogyx, and Enochian Biosciences, outside the submitted work. JD reports paid compensation for consulting for the Public Health Company and Allen & Co. MJP reports consulting fees from Gilead Sciences and AstraZeneca, outside the submitted work. All other authors report no potential conflicts.

## FIGURE LEGENDS

**Supplemental Table 1: Characteristics of study participants**.

Characteristics of all study participants, stratified by Long COVID status.

**Supplemental Figure 1: PhIP-Seq is unable to distinguish Long COVID symptom phenotypes from controls**.

(A) Logistic regression receiver operating characteristic (ROC) curves for Long COVID patients with different specified symptom phenotypes relative to patients previously infected with COVID without the phenotype, or (B) with and without EBV laboratory findings.

## Notes

### Summary of Updates

1. Update to contact information for corresponding author. 2. Added Supplemental Table 1, which had been omitted in error from the initial submission.

## REFERENCES

1. Nalbandian A et al. Post-acute COVID-19 syndrome [Internet]. Nat. Med. [published online ahead of print: March 22, 2021]; doi:10.1038/s41591-021-01283-z

2. Davis HE, McCorkell L, Vogel JM, Topol EJ. Long COVID: major findings, mechanisms and recommendations. Nat. Rev. Microbiol. 2023;1–14.

3. Peluso MJ, Deeks SG. Early clues regarding the pathogenesis of long-COVID. Trends Immunol. 2022;43(4):268–270.

4. Proal AD, VanElzakker MB. Long COVID or Post-acute Sequelae of COVID-19 (PASC): An Overview of Biological Factors That May Contribute to Persistent Symptoms [Internet]. Frontiers in Microbiology 2021;12. doi:10.3389/fmicb.2021.698169

5. Peluso MJ, Donatelli J, Henrich TJ. Long-term immunologic effects of SARS-CoV-2 infection: leveraging translational research methodology to address emerging questions. Transl. Res. 2022;241:1–12.

6. Zuo Y et al. Prothrombotic autoantibodies in serum from patients hospitalized with COVID-19 [Internet]. Sci. Transl. Med. 2020;12(570). doi:10.1126/scitranslmed.abd3876

7. Chang SE et al. New-onset IgG autoantibodies in hospitalized patients with COVID-19. Nat. Commun. 2021;12(1):5417.

8. Woodruff MC et al. Dysregulated naïve B cells and de novo autoreactivity in severe COVID-19 [Internet]. Nature [published online ahead of print: August 31, 2022]; doi:10.1038/s41586-022-05273-0

9. Bastard P et al. Autoantibodies against type I IFNs in patients with life-threatening COVID-19 [Internet]. Science 2020;370(6515). doi:10.1126/science.abd4585

10. Bastard P et al. Autoantibodies neutralizing type I IFNs are present in ∼4% of uninfected individuals over 70 years old and account for ∼20% of COVID-19 deaths [Internet]. Sci Immunol 2021;6(62). doi:10.1126/sciimmunol.abl4340

11. Vazquez SE et al. Neutralizing Autoantibodies to Type I Interferons in COVID-19 Convalescent Donor Plasma. J. Clin. Immunol. 2021;41(6):1169–1171.

12. van der Wijst MGP et al. Type I interferon autoantibodies are associated with systemic immune alterations in patients with COVID-19. Sci. Transl. Med. 2021;13(612):eabh2624.

13. Son K et al. Circulating anti-nuclear autoantibodies in COVID-19 survivors predict long-COVID symptoms [Internet]. Eur. Respir. J. [published online ahead of print: September 22, 2022]; doi:10.1183/13993003.00970-2022

14. Seeßle J et al. Persistent symptoms in adult patients one year after COVID-19: a prospective cohort study [Internet]. Clin. Infect. Dis. [published online ahead of print: July 5, 2021]; doi:10.1093/cid/ciab611

15. Su Y et al. Multiple Early Factors Anticipate Post-Acute COVID-19 Sequelae [Internet]. Cell 2022;0(0). doi:10.1016/j.cell.2022.01.014

16. Peluso MJ, Thomas IJ, Munter SE, Deeks SG, Henrich TJ. Lack of Antinuclear Antibodies in Convalescent Coronavirus Disease 2019 Patients With Persistent Symptoms. Clin. Infect. Dis. 2022;74(11):2083–2084.

17. Schultheiß C et al. The IL-1β, IL-6, and TNF cytokine triad is associated with post-acute sequelae of COVID-19. Cell Rep Med 2022;3(6):100663.

18. Klein J et al. Distinguishing features of Long COVID identified through immune profiling [Internet]. medRxiv [published online ahead of print: August 10, 2022]; doi:10.1101/2022.08.09.22278592

19. Bodansky A et al. NFKB2 haploinsufficiency identified via screening for IFNα2 autoantibodies in children and adolescents hospitalized with SARS-CoV-2-related complications [Internet]. J. Allergy Clin. Immunol. [published online ahead of print: December 9, 2022]; doi:10.1016/j.jaci.2022.11.020

20. Mandel-Brehm C et al. Kelch-like Protein 11 Antibodies in Seminoma-Associated Paraneoplastic Encephalitis. N. Engl. J. Med. 2019;381(1):47–54.

21. O’Donovan B et al. High-resolution epitope mapping of anti-Hu and anti-Yo autoimmunity by programmable phage display. Brain Commun 2020;2(2):fcaa059.

22. Vazquez SE et al. Identification of novel, clinically correlated autoantigens in the monogenic autoimmune syndrome APS1 by proteome-wide PhIP-Seq [Internet]. Elife 2020;9. doi:10.7554/eLife.55053

23. Vazquez SE et al. Autoantibody discovery across monogenic, acquired, and COVID-19-associated autoimmunity with scalable PhIP-seq [Internet]. Elife 2022;11. doi:10.7554/eLife.78550

24. Larman HB et al. Autoantigen discovery with a synthetic human peptidome. Nat. Biotechnol. 2011;29(6):535–541.

25. Peluso MJ et al. Persistence, magnitude, and patterns of postacute symptoms and quality of life following onset of SARS-CoV-2 infection: cohort description and approaches for measurement. In: Open forum infectious diseases. Oxford University Press US; 2022:ofab640

26. Peluso MJ et al. Impact of pre-existing chronic viral infection and reactivation on the development of long COVID [Internet]. J. Clin. Invest. [published online ahead of print: December 1, 2022]; doi:10.1172/JCI163669

