## Supplemental Table 1 for "Autoantigen profiling reveals a shared post-COVID signature in fully recovered and Long COVID patients"

Supplemental Table 1. Participant characteristics

|  | PASC | No PASC | Total |
| --- | --- | --- | --- |
|  | n=121 | n=64 | n=185 |
| Days post-symptom onset | 121 (107 to 129) | 121 (108 to 128) | 121 (108 to 129) |
| Number of COVID-attributed symptoms at time of visit | 4 (2 to 8) | 0 (0 to 0) | 2 (0 to 6) |
| Age in years | 46 (38 to 55) | 48 (38 to 58) | 48 (38 to 56) |
| Female birth sex | 57 (47.1%) | 24 (37.5%) | 81 (43.8%) |
| Race/ethnicity |  |  |  |
| Hispanic/Latino | 44 (37.0%) | 12 (19.4%) | 56 (30.9%) |
| White | 61 (51.3%) | 34 (54.8%) | 95 (52.5%) |
| Black/African American | 6 (5.0%) | 4 (6.5%) | 10 (5.5%) |
| Asian | 7 (5.9%) | 9 (14.5%) | 16 (8.8%) |
| Pacific Islander/Native Hawaiian | 1 (0.8%) | 3 (4.8%) | 4 (2.2%) |
| Hospitalized during COVID-19 illness | 30 (24.8%) | 13 (20.3%) | 43 (23.2%) |
| Medical Comorbidities |  |  |  |
| Autoimmune disease | 9 (7.4%) | 1 (1.6%) | 10 (5.4%) |
| Cancer treated within past 2 years | 4 (3.3%) | 1 (1.6%) | 5 (2.7%) |
| Diabetes | 11 (9.4%) | 8 (12.5%) | 19 (10.5%) |
| HIV | 31 (25.6%) | 8 (12.5%) | 39 (21.1%) |
| Heart attack or heart failure | 3 (2.5%) | 2 (3.1%) | 5 (2.7%) |
| Hypertension | 30 (25.0%) | 6 (9.4%) | 36 (19.6%) |
| Lung disease | 18 (15.0%) | 13 (20.3%) | 31 (16.8%) |
| Kidney disease | 1 (0.8%) | 1 (1.6%) | 2 (1.1%) |
| History of tobacco smoking | 37 (32.2%) | 15 (26.8%) | 52 (30.4%) |

All values are median (IQR) unless otherwise noted.
