## Supplementary figures and images for "Autoantigen profiling reveals a shared post-COVID signature in fully recovered and Long COVID patients"

### Supplemental Figure 1

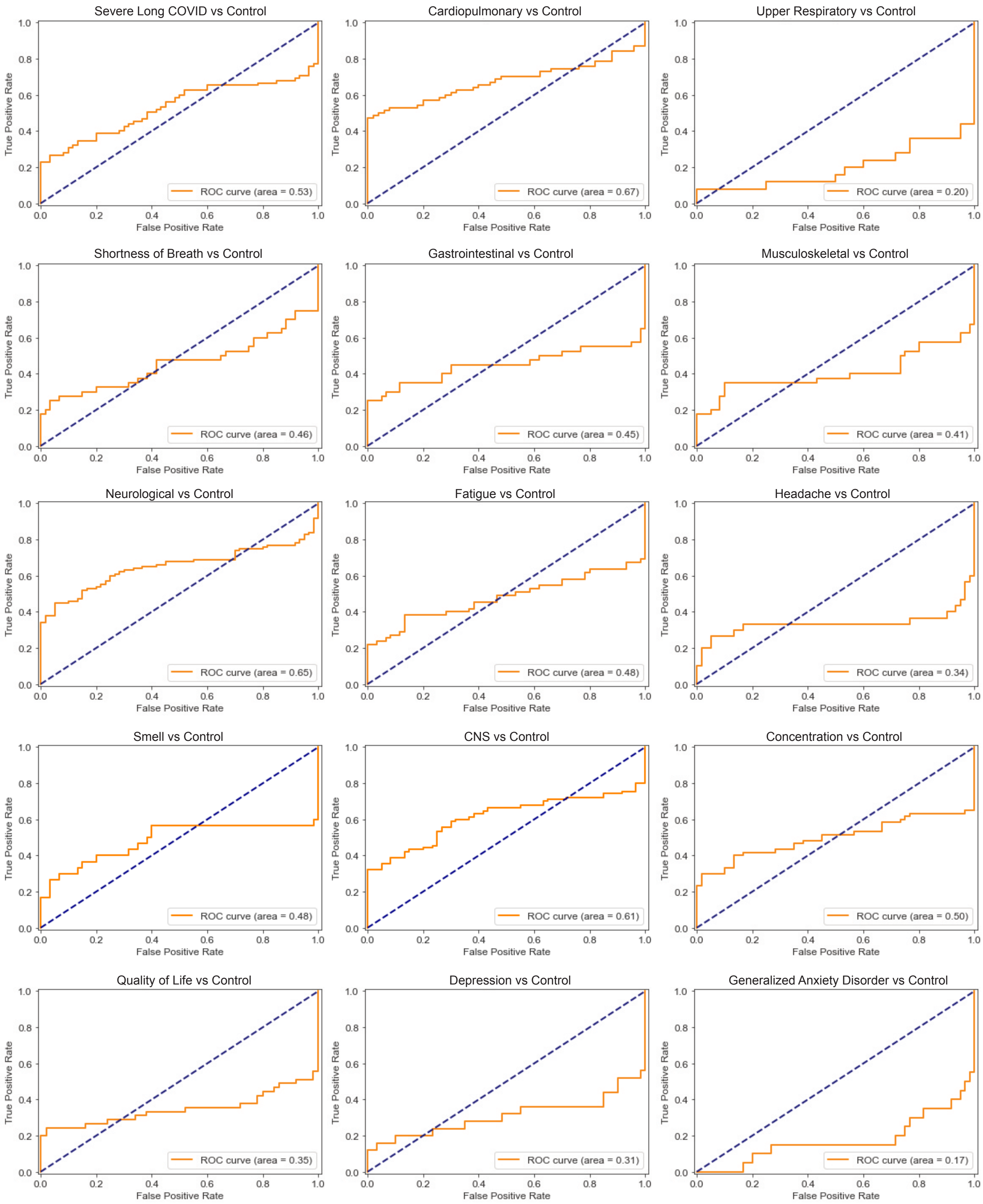

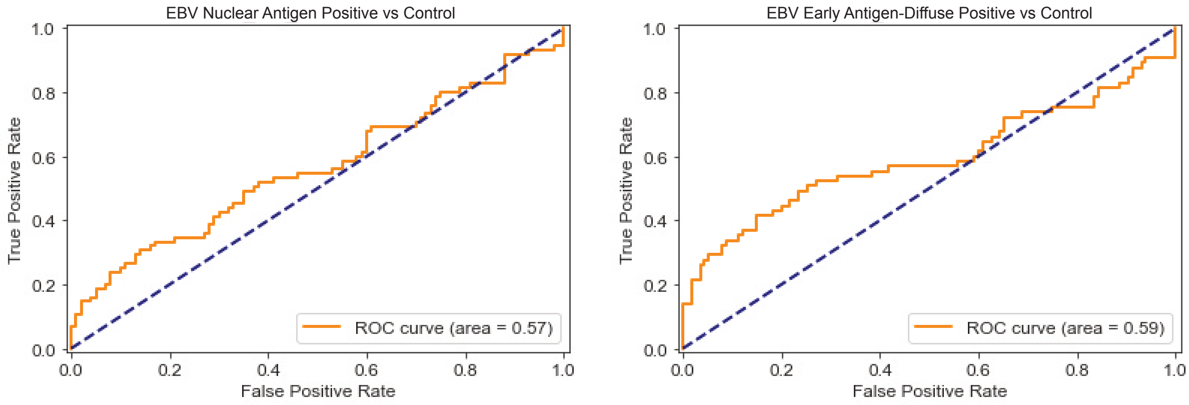
